# Distributed Counterfactual Modeling Approach for Investigating Hospital-Associated Racial Disparities in COVID-19 Mortality

**DOI:** 10.1101/2021.09.14.21263596

**Authors:** Mackenzie Edmondson, Chongliang Luo, Nazmul Islam, David Asch, Jiang Bian, Yong Chen

**Affiliations:** Department of Biostatistics, Epidemiology, and Informatics, University of Pennsylvania Perelman School of Medicine, Philadelphia, PA; OptumLabs at UnitedHealth Group, Minnetonka, MN; Division of General Internal Medicine, University of Pennsylvania, Philadelphia, PA; Leonard Davis Institute of Health Economics, University of Pennsylvania, Philadelphia, PA; Department of Health Outcomes and Biomedical Informatics, College of Medicine, University of Florida, Gainesville, FL; Cancer Informatics Shared Resource, University of Florida Health Cancer Center, Gainesville, FL

## Abstract

Several studies have found that black patients are more likely than white patients to test positive for or be hospitalized with COVID-19, but many of these same studies have found no difference in in-hospital mortality. These studies may have underestimated racial differences due to reliance on data from a single hospital system, as adequate control of patient-level characteristics requires aggregation of highly granular data from several institutions. Further, one factor thought to contribute to disparities in health outcomes by race is site of care. Several differences between black and white patient populations, such as access to care and referral patterns among clinicians, can lead to patients of different races largely attending different hospitals. We sought to develop a method that could study the potential association between attending hospital and racial disparity in mortality for COVID-19 patients without requiring patient-level data sharing among collaborating institutions. We propose a novel application of a distributed algorithm for generalized linear mixed modeling (GLMM) to perform counterfactual modeling and investigate the role of hospital in differences in COVID-19 mortality by race. Our counterfactual modeling approach uses simulation to randomly assign black patients to hospitals in the same distribution as those attended by white patients, quantifying the difference between observed mortality rates and simulated mortality risk following random hospital assignment. To illustrate our method, we perform a proof-of-concept analysis using data from four hospitals within the OneFlorida Clinical Research Consortium. Our approach can be used by investigators from several institutions to study the impact of admitting hospital on COVID-19 mortality, a critical step in addressing systemic racism in modern healthcare.

## Introduction

Throughout the COVID-19 pandemic, several studies have found that black patients are more likely than white patients to test positive for or be hospitalized with COVID-19 [1-5]. Many of these studies also found that there was no difference in in-hospital mortality for black and white patients after adjusting for patient-level sociodemographic and clinical characteristics [1-2, 4-5]. However, those studies may have underestimated racial differences because they relied on data from single hospital systems, which might have more homogeneous and less broadly representative populations. A much larger study reflecting data from approximately 1,200 US hospitals found residual racial differences [10]. That study was possible because of a single data source from a large insurance company, but many questions cannot be answered without data that are already aggregated and in a sufficiently granular level.

The challenges of aggregating highly granular data are critically relevant to investigations of racial disparities in health care because of the frequent need to control for patient-level characteristics. Those statistical adjustments often reveal mediating pathways because individual characteristics that seemingly confound associations by race might themselves be the products of past racial injustice. For example, making statistical adjustments for insurance type, area of residence, and comorbidities might seem to allow estimation of isolated effects of race, but if those factors are themselves the products of racial discrimination those adjustments can risk obscuring racial differences in health outcomes rather than identifying them.

Indeed, one factor thought to contribute to disparities in health outcomes by race is site of care [6]. Patients of different races tend to live in different areas and so their sources of care and referral patterns tend to differ. For example, a study examining hospital-level racial disparities in acute myocardial infarction treatment and outcomes found that black patients tend to receive care at different hospitals than white patients [7]. Another study found that black and white patients tend to be treated by different physicians, with physicians for black patients possibly less qualified and lacking access to critical clinical resources relative to physicians who tend to treat white patients [8]. These reported differences are likely associated with a number of differences in the black and white patient populations, including access to care, referral patterns among clinicians, and susceptibility to harmful social and environmental exposures. Repeatedly, racial differences in health outcomes have been attributed to differences in care site.

To investigate the potential association between admitting hospital and racial disparity in mortality for COVID-19 patients, a counterfactual modeling approach can be used [9]. If we assume that black and white patients each attend hospitals according to some underlying distribution, then using counterfactual modeling, we can simulate hospital assignments for black patients under the hypothetical scenario that they attend hospitals in the same distribution as white patients. Assuming the existence of hospital-level effects affecting in-hospital mortality, one may hypothesize that simulated mortality rate under this scenario would be lower than observed mortality rate for black patients if one assumes hospitals with predominately white patients have higher quality of care on average than hospitals with mostly black patients. This counterfactual modeling approach can aid in unveiling disparities in health outcomes associated with site of care, a crucial first step in addressing systemic racism in modern healthcare.

A recent study by Asch et al. (2020) explored the potential connection between admitting hospital and racial disparities in COVID-19 using counterfactual modeling, fitting a generalized linear mixed model (GLMM) to model log odds of mortality while adjusting for both common patient-level fixed effects as well as hospital-specific fixed and random effects [10]. Estimation of hospital-specific effects is the key component for counterfactual modeling, allowing for estimating patient-specific mortality risk as if the patient (counterfactually) attended a hospital different from the one they truly attended. While effective for this particular study, which featured a centralized data repository allowing direct access to all patient data, GLMM is not able to be used if data are not centralized. Patient-level data sharing among hospitals is often not possible due to regulations protecting patient privacy. If each participating institution can instead share only aggregate data, a method allowing for distributed estimation of GLMM is necessary.

In this work, we propose a novel application of a recently developed algorithm for performing GLMM estimation [11] to study hospital-associated racial disparity in COVID-19 mortality via counterfactual modeling. We refer to this approach as the distributed penalized quasi-likelihood (dPQL) algorithm. The dPQL is based on the penalized quasi-likelihood (PQL) algorithm, an iterative procedure for GLMM estimation [12-13] that has commonly been used to fit GLMM due to its ease of computation; it usually achieves convergence in several iterations. The dPQL algorithm is a distributed version of PQL, where in each iteration it requires participating hospitals to share aggregate, summary-level data rather than patient-level data. The result of the dPQL algorithm is lossless, as fixed-effect and random-effect estimates are identical to those produced by traditional PQL as if one had access to centralized patient-level data. After estimating the fixed and random effects via the distributed algorithm, individual hospitals within a multi-site study can further estimate counterfactual mortality risk for each of their own patients, quantifying the effect of their patients receiving care at another hospital in the study. Counterfactual mortality risk, as will be demonstrated, can also be obtained in a distributed manner (without sharing patient-level data). Thus, the above distributed analysis framework creates the potential for large-scale, multi-site studies for assessing hospital-associated racial disparities when sharing patient-level data is not possible.

The remainder of this work is structured as follows. In the Methods section, we first provide an overview of how to estimate patient-specific mortality risk at a given hospital and propose using the dPQL algorithm for these purposes. We then describe estimation of counterfactual mortality risks for each patient within each hospital before outlining a simulation approach which can be used to investigate hospital-associated racial disparities in COVID-19 mortality vis counterfactual modeling. Lastly, we offer an illustrative example of how our proposed application of the dPQL algorithm can be used in practice, using it to analyze patient data from four hospitals within the OneFlorida Clinical Research Consortium. In our analysis, we sought to determine whether observed 30-day in-hospital mortality rate for Non-Hispanic Black (NHB) patients differed from simulated overall mortality risk, calculated under the hypothetical scenario that NHB patients attended hospitals in the same distribution as Non-Hispanic White (NHW) patients. We present results for our proof-of-concept simulation before concluding with a discussion on the implications of our work.

## Methods

### Modeling Mortality Risk: Generalized Linear Mixed Model

Investigating the association between attending hospital and racial disparities in COVID-19 mortality via counterfactual modeling requires a statistical modeling approach that accounts for hospital-specific effects and is compatible with the data sharing arrangement in place among participating hospitals. Suppose *H* hospitals, each with *n*_*h*_ patients, are willing to participate in a multi-site analysis. Each hospital has direct access to its own relevant patient-level or hospital-level covariates ***X***_***ih***_ for each patient *i* at hospital 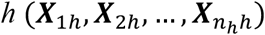 and outcome of interest *y*_*ih*_ for each patien 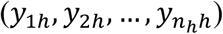 In the context of our study question, suppose this outcome is binary. If hospitals agree to share patient-level data across hospitals, allowing for a centralized data repository, a generalized linear mixed modeling (GLMM) approach can be implemented using the pooled patient-level data. The mean and variance using GLMM are specified as

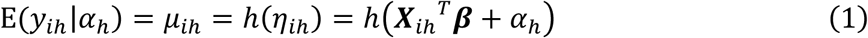

and

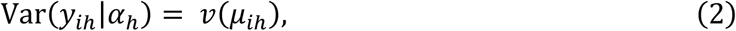

where *g* = *h*^-1^ is the link function that connects the conditional patient-level means *μ*_*ij*_ to the linear predictor *η*_*ij*_ and *v*(·) is the variance function. The random effects *α*_*h*_ are assumed to follow a normal distribution with mean 0 and variance θ. In our context, the GLMM method allows for modeling common fixed effects ***β*** (*β*_1_, *β*_*2*_, …, *β*_*p*_) for *p* covariates at either the patient or hospital level as well as hospital-specific random effects ***α***, such as a random intercept estimated for each hospital (*α*_1_, *α*_*2*_, …, *α*_*H*_). In our modeling below, hospital-specific random intercepts are the only random effects modeled. Incorporating additional hospital-specific random effects is possible if desired. Estimated fixed and random effects, 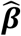 and 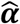, respectively, can then be used to estimate mortality risk for each patient in the sample.

### Distributed Penalized Quasi-Likelihood (dPQL) algorithm

In settings where patient-level data cannot be shared, collaborating hospitals in a multi-site study often agree to conduct a distributed data analysis. When analyzing data distributively, patient-level data remain within institution, with each hospital analyzing its own data directly before sending aggregate results to a coordinating center. Available methods for performing distributed data analysis vary in their accuracy relative to pooled analysis methods and communication required among participating institutions to obtain results. Most distributed analysis methods require a lead site to coordinate communication among the collaborating sites and aggregate hospital-specific results.

We propose using a recently developed distributed penalized quasi-likelihood (dPQL) algorithm [11] for performing distributed GLMM. Estimation via penalized quasi-likelihood (PQL) is one of several possible approaches for fitting a GLMM. The PQL algorithm iteratively fits the linear mixed model (LMM)

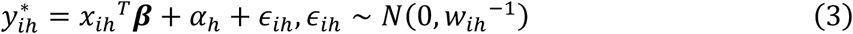

with the working outcome

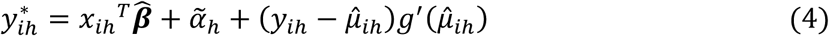

and the weight

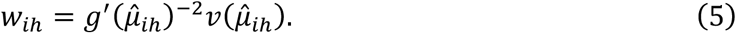

The obtained effect and variance estimates are denoted as 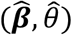. The dPQL algorithm is a distributed version of PQL, where in each iteration participating hospitals are required to share only aggregate data with the lead site to fit the LMM. This is done by using a distributed linear mixed model (DLMM) algorithm [14]. Since the DLMM algorithm is lossless, resulting in estimates identical to those produced by the LMM, the dPQL algorithm also obtains the same estimates as those obtained by the PQL algorithm as if all patient-level data are available. The dPQL algorithm is summarized below.

#### The dPQL algorithm

1. Initialization: The lead site sends an initial value for the fixed effects ***β***^(0)^ and random effects ***α***^(0)^ = 0 to all participating sites *h* = 1, *2*, …, *H*(including the lead site).
2. For iteration *s* = 0, 1, …, *S* and within each site *h* = 1, *2*, …, *H*:
  2.1. Site *h* calculates the working outcome 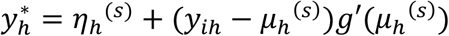, linear predictor *η*_*h*_^(*s*)^ = *X*_*h*_*β*^(*s*)^ + *α*_*h*_^(*s*)^, and weights *W*_*h*_ = *diag*{*g*′(*μ*_*h*_^(*s*)^)^-*2*^*v*(*μ*_*h*_^(*s*)^)}.
  2.2. Site *h* calculates the following aggregated data, communicating each measure to the lead site:
    - *p* × *p* matrix: 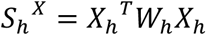
    - *p*-dimensional vector: 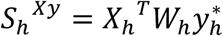
    - scalars: 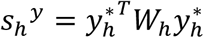 and sample size *n*_*h*_
  2.3. Lead site fits weighted DLMM algorithm based on aggregated data from 2.2. to obtain updated ***β***^(*s*+1)^ and ***α***^(*s*+*2*)^, returning these to the collaborating sites to replace initial values.
3. 2.1. through 2.3. are repeated until convergence, e.g. || *η*^(*s*+1)^ − *η*^(*s*)^‖ / ‖ *η*^(*s*)^ ‖ < 1e-6. The final estimates are 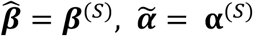 and 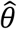, all obtained in the final iteration *S*.

### Distributed Counterfactual Modeling

Given the distributively estimated fixed effect 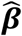 and random effects 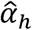 for hospital h, for patient *i* who attended hospital *h*, their estimated mortality risk can be calculated as

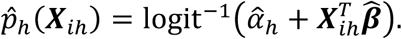

We refer to this as the factual mortality risk for patient *i*. Our primary interest for this analysis is to investigate the association between attending hospital and in-hospital mortality differences for Non-Hispanic Black (NHB) and Non-Hispanic White (NHW) patients. To do this, we use a counterfactual modeling approach to study whether the observed in-hospital mortality rate for NHB patients differs from their simulated in-hospital mortality rate given they (hypothetically) attended hospitals in the same distribution as NHW patients [9]. Counterfactual mortality risk estimates differ from the factual mortality risk estimates defined above in that they are purely hypothetical, with estimates calculated using an estimated random intercept for a hospital that a given patient did not truly attend. The difference between factual and counterfactual mortality risk estimates and how they are calculated are detailed in Figure 1. While 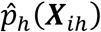 denotes the factual mortality risk estimate for patient *i* at hospital *h*, we can calculate counterfactual mortality risk estimates for all patients as if they attended any of the hospitals participating in the study, denoting counterfactual mortality estimates using 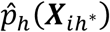 where *h* ≠ *h*^*^. Using this notation, *h* denotes the hypothetical attending hospital, the hospital contributing the estimated random intercept, while *h*^*^ denotes the hospital that patient truly attended.

**Figure 1.**
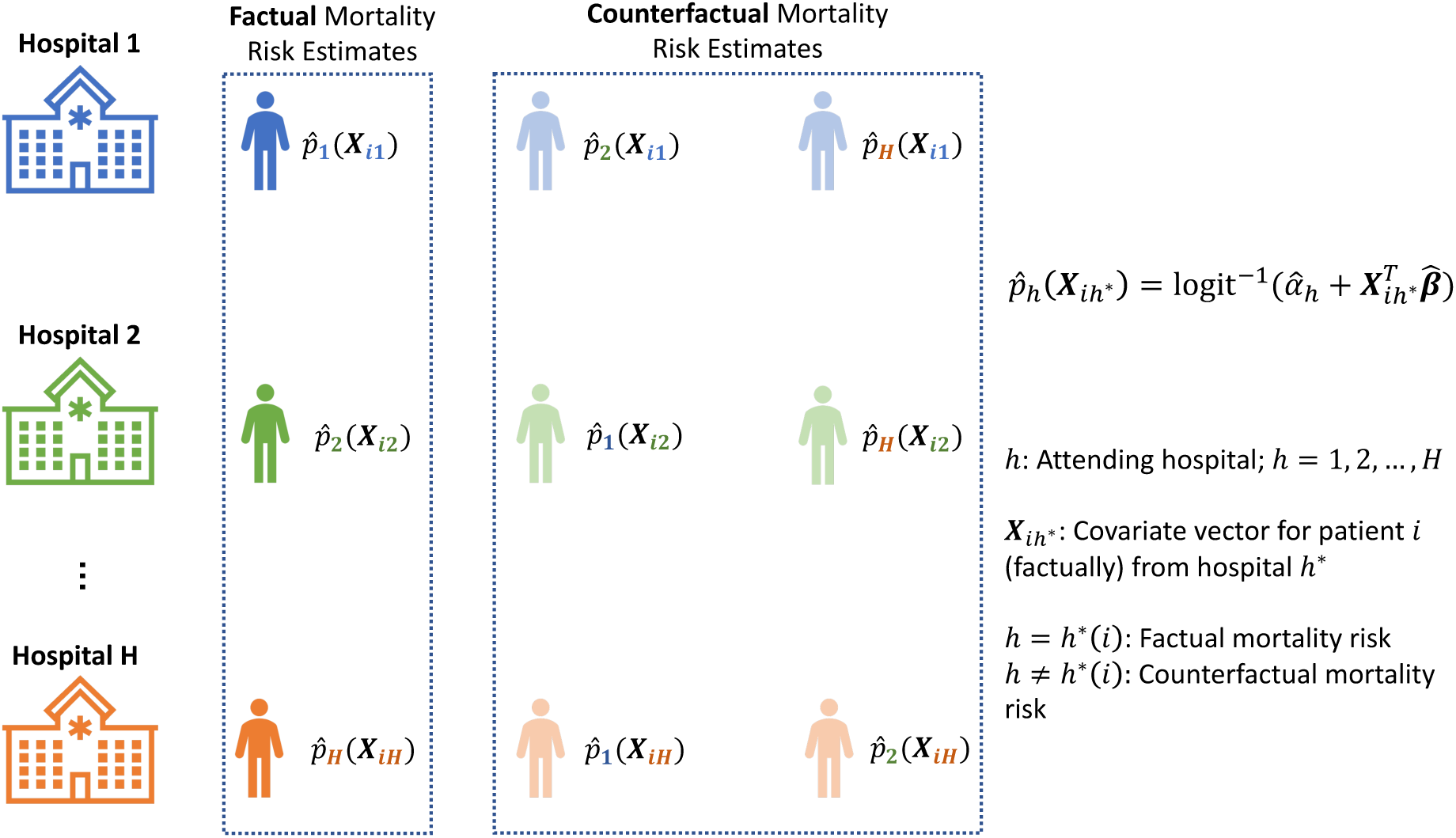
Schematic diagram demonstrating how to calculate both factual and counterfactual mortality risk for patients as if they (hypothetically) attended each hospital. Each row depicts how patient data within a given hospital can be used to estimate mortality risks as if their patients had attended any hospital in the study. The first column features factual mortality risk estimates, calculated using common fixed effect estimates, the random intercept estimate for that given hospital, and the patient data for that hospital. The last two columns provide examples for computable counterfactual mortality risks, which use random intercept estimates for hospitals different from those that patients truly attended.

We next quantify racial disparity by conducting a simulation to investigate the counterfactual mortality rate of black patients had they been admitted to hospitals in the same distribution as white patients while retaining their sociodemographic and clinical characteristics.

Our simulation procedure is depicted at a high level in Figure 2. Suppose the number of black patients at each hospital *h, h* = 1, …, *H*, is *n*_*hB*_. Each hospital shares the number of white patients at their respective hospitals, *n*_*hW*_ = *n*_1*W*_, *n*_*2W*_, …, *n*_*HW*_, with the coordinating center. These are used to determine the relative proportion of total white patients at each hospital, *ŵ*_*h*_ = *ŵ*_1_,*ŵ*_2_, …, *ŵ*_*H*_, which are communicated to each hospital. Next, for replicate *s* = 1, …, *S* of the simulation within hospital *h*, a multinomial distribution with probabilities equal to *ŵ*_1_,*ŵ*_2_, …, *ŵ*_*H*_, is used to assign each black patient to a hospital in the study. Assume patient *i* is assigned to hospital *h*^*^. Hospital assignments may be factual if *h* = *h*^*^ or counterfactual if *h* ≠ *h*^*^. Using these hospital assignments, (counter)factual mortality risk estimates are calculated for each patient *i* as 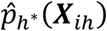.

**Figure 2.**
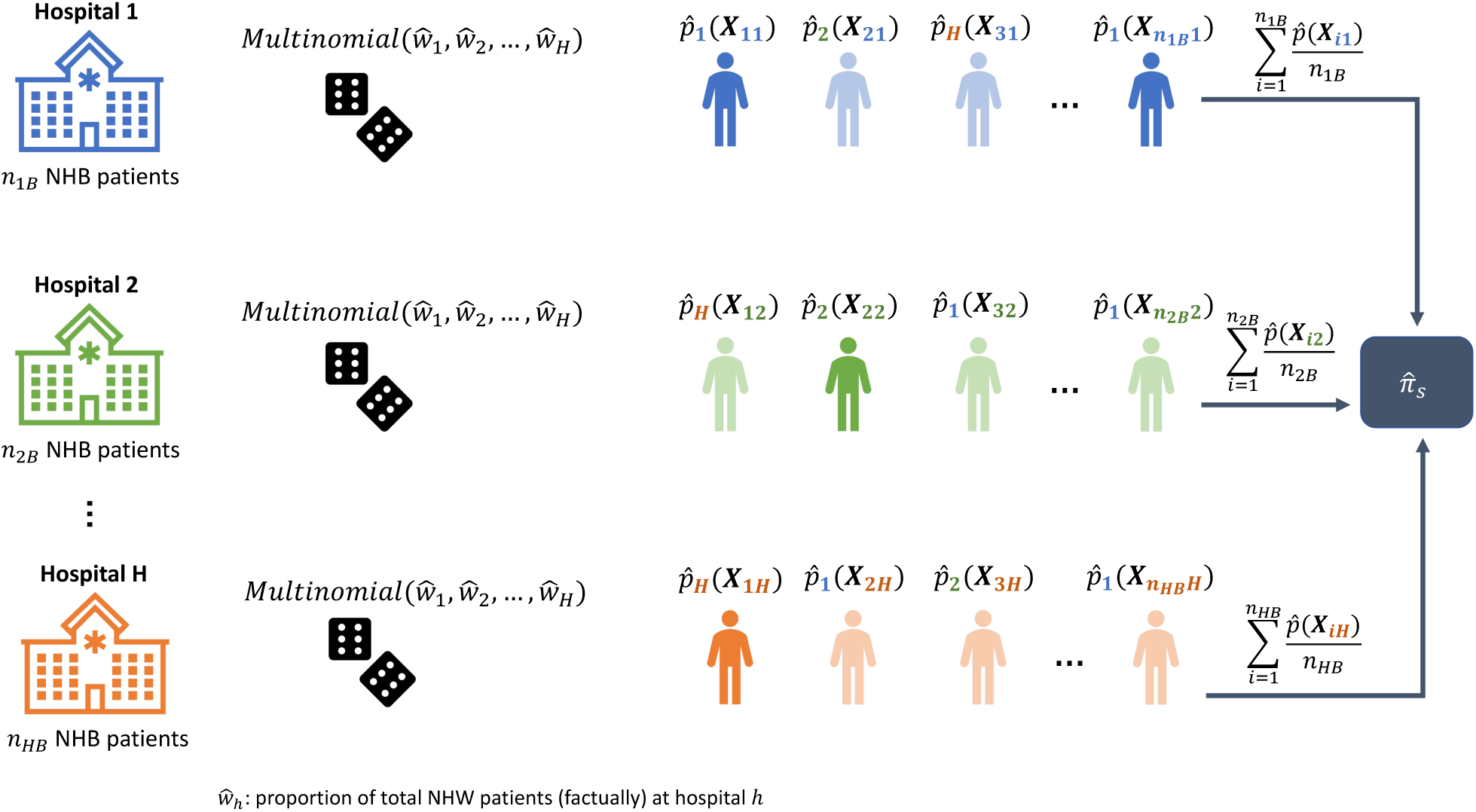
Schematic overview of our simulation procedure for calculating simulated mortality risk estimates using counterfactual modeling. In each iteration of the simulation, within each hospital, a multinomial distribution with probabilities equal to the proportion of total white patients at each hospital (*ŵ*_1_,*ŵ*_2_, …, *ŵ*_*H*_) is used to assign each black patient to a hospital. Hospital assignments can be factual (denoted by darker colors) or counterfactual (denoted by lighter colors). Hospital assignments are then used to calculate mortality risk estimates for each black patient as if they attended the assigned hospital. Refer to Figure 1 to see how mortality risk estimates are calculated. Each hospital then averages their patient-level mortality risk estimates and communicates them to the coordinating center, where overall simulated mortality risk estimate 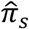 is obtained for each simulation replicate. Multinomial hospital assignment and resultant calculation of (counter)factual mortality risk estimates is performed in each simulation replicate.

To preserve patient privacy, individual patient mortality risk estimates are averaged within hospital *h* as

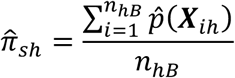

and communicated to the coordinating site. Simulated overall mortality risk 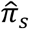 is then calculated using the following:

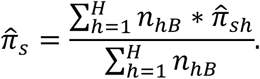

Since we are interested in whether hypothetical hospital assignment affects mortality risk for black patients, in each iteration of the simulation, we calculate the difference between observed black patient mortality rate 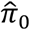 and simulated black patient mortality risk 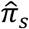. The mean difference across iterations can be reported along with an empirical 95% confidence interval (using the 2.5^th^ and 97.5^th^ percentiles of 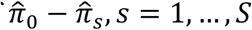) to quantify uncertainty in the resulting difference.

### Illustrative Example Using Multi-Hospital Real-World Data

We illustrate our application of the dPQL algorithm to study hospital-associated racial disparities in COVID-19 mortality using data from the OneFlorida Clinical Research Consortium, a centralized data repository comprising data for over 74% of Floridians.

OneFlorida data include records from Medicaid and Medicare claims, cancer registries, and electronic health records from various clinical partners in the state of Florida. As a proof-of-concept analysis, we conducted a counterfactual modeling simulation using patient data from four hospitals in the OneFlorida Clinical Research Consortium. Patients included in the analysis were required to be hospitalized with COVID-19 and have index hospitalization dates between 3/1/20 and 2/28/21 to ensure 30 days of follow up for each patient. Using the dPQL algorithm, we distributively modeled the log odds of 30-day COVID-19 in-hospital mortality as a function of various patient characteristics, including age, gender, a collection of nine comorbidities, and index quarter, defined as one of four three-month intervals when a given patient was admitted. Definitions for each quarter, as well as further details concerning the included comorbidities and descriptive statistics for each hospital, are presented in Table 1.

**Table 1.** Summary of patient-level characteristics for patients hospitalized with COVID-19 at four hospitals within the OneFlorida Clinical Research Consortium.

|  | **Hospital A**<br>**(N=2232)** | **Hospital B**<br>**(N=1915)** | **Hospital C**<br>**(N=1047)** | **Hospital D**<br>**(N=565)** | **Total**<br>**(N=5759)** |
| --- | --- | --- | --- | --- | --- |
| **Age** |  |  |  |  |  |
| [18,65) | 1005 (45.0%) | 1111 (58.0%) | 566 (54.1%) | 198 (35.0%) | 2880 (50.0%) |
| ≥ 65 | 1227 (55.0%) | 804 (42.0%) | 481 (45.9%) | 367 (65.0%) | 2879 (50.0%) |
| **Gender** |  |  |  |  |  |
| Female | 1249 (56.0%) | 1082 (56.5%) | 574 (54.8%) | 313 (55.4%) | 3218 (55.9%) |
| Male | 983 (44.0%) | 833 (43.5%) | 473 (45.2%) | 252 (44.6%) | 2541 (44.1%) |
| **Race** |  |  |  |  |  |
| Non-Hispanic White | 1432 (64.2%) | 892 (46.6%) | 485 (46.3%) | 331 (58.6%) | 3140 (54.5%) |
| Non-Hispanic Black | 800 (35.8%) | 1023 (53.4%) | 562 (53.7%) | 234 (41.4%) | 2619 (45.5%) |
| **Congestive Heart Failure** |  |  |  |  |  |
| Yes | 347 (15.5%) | 428 (22.3%) | 195 (18.6%) | 138 (24.4%) | 1108 (19.2%) |
| No | 1885 (84.5%) | 1487 (77.7%) | 852 (81.4%) | 427 (75.6%) | 4651 (80.8%) |
| **Myocardial Infarction** |  |  |  |  |  |
| Yes | 109 (4.9%) | 184 (9.6%) | 66.0 (6.3%) | 56.0 (9.9%) | 415 (7.2%) |
| No | 2123 (95.1%) | 1731 (90.4%) | 981 (93.7%) | 509 (90.1%) | 5344 (92.8%) |
| **Peripheral Vascular Disease** |  |  |  |  |  |
| Yes | 223 (10.0%) | 344 (18.0%) | 134 (12.8%) | 86.0 (15.2%) | 787 (13.7%) |
| No | 2009 (90.0%) | 1571 (82.0%) | 913 (87.2%) | 479 (84.8%) | 4972 (86.3%) |
| **Cerebrovascular Disease** |  |  |  |  |  |
| Yes | 197 (8.8%) | 255 (13.3%) | 100 (9.6%) | 56.0 (9.9%) | 608 (10.6%) |
| No | 2035 (91.2%) | 1660 (86.7%) | 947 (90.4%) | 509 (90.1%) | 5151 (89.4%) |
| **Dementia** |  |  |  |  |  |
| Yes | 239 (10.7%) | 125 (6.5%) | 99.0 (9.5%) | 54.0 (9.6%) | 517 (9.0%) |
| No | 1993 (89.3%) | 1790 (93.5%) | 948 (90.5%) | 511 (90.4%) | 5242 (91.0%) |
| **Chronic Pulmonary Disease** |  |  |  |  |  |
| Yes | 480 (21.5%) | 538 (28.1%) | 215 (20.5%) | 149 (26.4%) | 1382 (24.0%) |
| No | 1752 (78.5%) | 1377 (71.9%) | 832 (79.5%) | 416 (73.6%) | 4377 (76.0%) |
| **Diabetes** |  |  |  |  |  |
| Yes | 734 (32.9%) | 682 (35.6%) | 307 (29.3%) | 222 (39.3%) | 1945 (33.8%) |
| No | 1498 (67.1%) | 1233 (64.4%) | 740 (70.7%) | 343 (60.7%) | 3814 (66.2%) |
| **Diabetes w/ Chronic Complication** |  |  |  |  |  |
| Yes | 350 (15.7%) | 437 (22.8%) | 192 (18.3%) | 128 (22.7%) | 1107 (19.2%) |
| No | 1882 (84.3%) | 1478 (77.2%) | 855 (81.7%) | 437 (77.3%) | 4652 (80.8%) |
| **Any Malignant Tumor** |  |  |  |  |  |
| Yes | 161 (7.2%) | 183 (9.6%) | 94.0 (9.0%) | 46.0 (8.1%) | 484 (8.4%) |
| No | 2071 (92.8%) | 1732 (90.4%) | 953 (91.0%) | 519 (91.9%) | 5275 (91.6%) |
| **Index Quarter** |  |  |  |  |  |
| Quarter 1 (3/20 - 5/20) | 123 (5.5%) | 182 (9.5%) | 59.0 (5.6%) | 20.0 (3.5%) | 384 (6.7%) |
| Quarter 2 (6/20 - 8/20) | 861 (38.6%) | 711 (37.1%) | 504 (48.1%) | 166 (29.4%) | 2242 (38.9%) |
| Quarter 3 (9/20 - 11/20) | 444 (19.9%) | 268 (14.0%) | 164 (15.7%) | 97.0 (17.2%) | 973 (16.9%) |
| Quarter 4 (12/20 - 2/21) | 804 (36.0%) | 754 (39.4%) | 320 (30.6%) | 282 (49.9%) | 2160 (37.5%) |
| **30-Day In-Hospital Mortality** |  |  |  |  |  |
| Deceased | 168 (7.5%) | 211 (11.0%) | 59.0 (5.6%) | 119 (21.1%) | 557 (9.7%) |
| Alive | 2064 (92.5%) | 1704 (89.0%) | 988 (94.4%) | 446 (78.9%) | 5202 (90.3%) |

Our primary analysis in this proof-of-concept study concerned calculating the difference between observed mortality rates and average simulated mortality risk estimates for all non-Hispanic black (NHB) patients across the four hospitals. We also performed a series of secondary analyses stratified by index quarter; four sub-analyses were conducted, with each only including patients with an index date for COVID-19 hospitalization in a particular quarter. Due to the rapid changes in the COVID-19 landscape as the pandemic progressed, including potential improvements in patient care and increasing prevalence of rare variants, we were interested in exploring whether the difference between observed and simulated mortality varied by quarter and if a trend in either direction was evident.

In Figure 3, we depict the relative distributions of non-Hispanic white (NHW) and NHB patients across the four hospitals, both within specific quarters and overall. Overall and in each quarter except for Quarter 1, Hospital A was the most well-attended, with the percentage of NHW patients attending Hospital A ranging from 38% in Quarter 1 to 52.5% in Quarter 3. For NHB patients, Hospital B was attended most overall and in each quarter except for Quarter 3. The percentage of NHB patients attending Hospital B ranged from 31.7% in Quarter 3 to 49.7% in Quarter 1. While NHW and NHB patients differed in this respect, the distributions of hospitals attended for each set of patients was largely similar. Distributions of hospitals attended across quarters were also similar, indicating that the racial makeup of the patient population at each hospital remained relatively constant throughout the twelve months of the COVID-19 pandemic studied in this analysis (March 2020 to February 2021).

**Figure 3.**
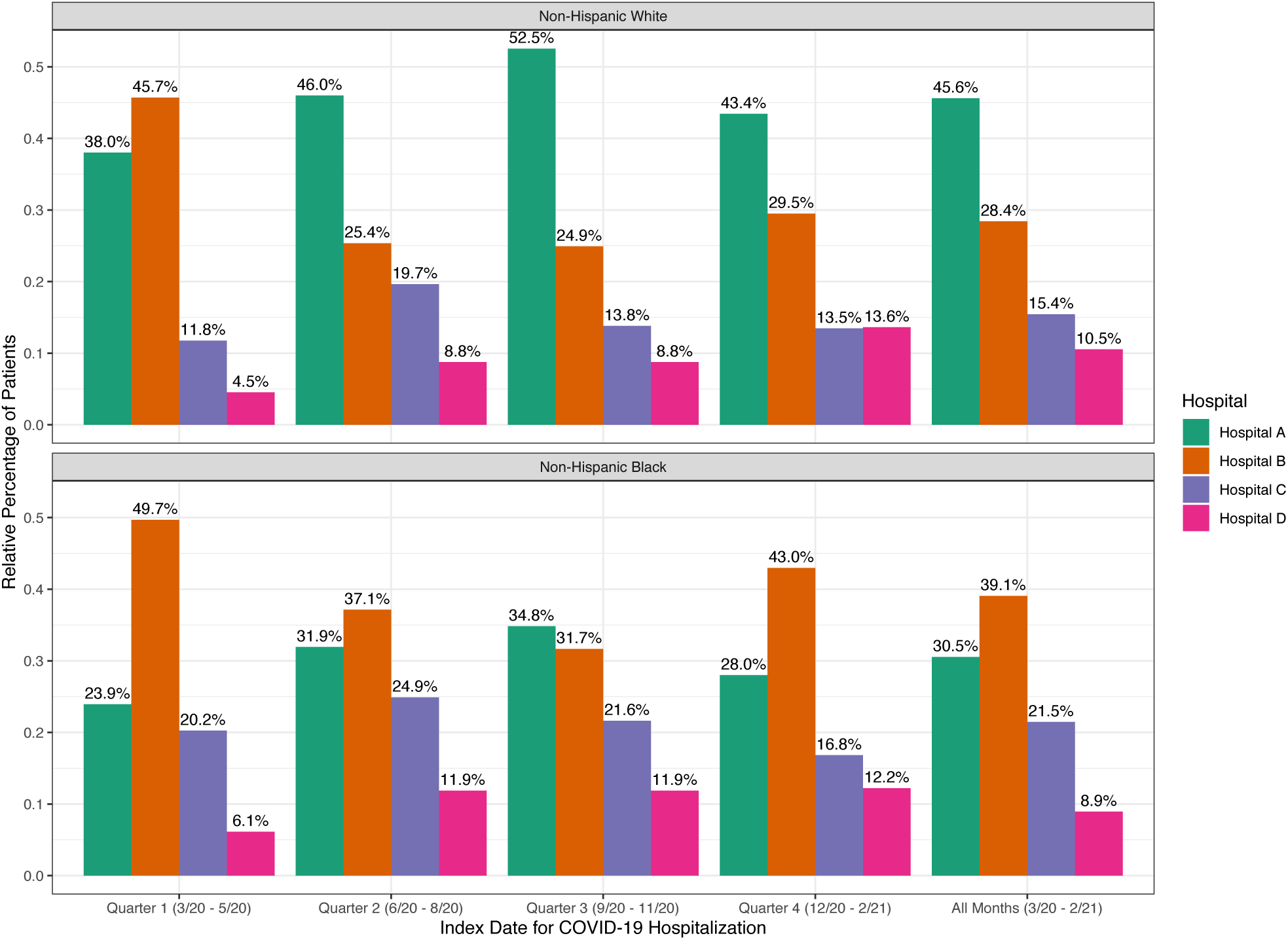
Bar plots depicting relative distributions of Non-Hispanic White (NHW) and Non-Hispanic Black (NHB) patients across hospitals, both within each index quarter and overall. Percentages above each bar represent the relative percentage of the total number of either NHW or NHB patients who attended each hospital. Dates are in the format “month/year”.

To conduct our simulation for both our primary and secondary (stratified) analyses, we first obtained the proportions of total NHW patients at each of the four hospitals, depicted in Figure 3. Then, within each hospital, we randomly assigned patients to one of the four hospitals using a multinomial distribution with probabilities equal to the four proportions found above. Simulated hospital assignments were used to calculate patient- and hospital-specific (counter)factual mortality risk estimates. Since OneFlorida data are centralized, we were able to average all mortality risk estimates from each hospital to calculate 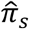. without needing to communicate risk estimate averages from each hospital to a coordinating center. However, if data were not centralized, identical results could be obtained by following the procedure outlined in the previous subsection. We conducted *S* = 500 iterations of the simulation for each of the five analyses, reporting the average difference in observed mortality rate and mean simulated mortality risk across iterations along with a corresponding 95% empirical confidence interval.

## Results

Boxplots summarizing results from each of our simulation studies are presented in Figure 4. Recall that the results presented here are intended to be the product of a proof-of-concept analysis to demonstrate the utility of this simulation method for performing counterfactual modeling. These results therefore should not be interpreted as having clinical significance. Using the complete set of patient data across all index dates, the mean difference across 500 iterations between observed in-hospital mortality rate 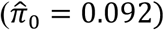 and simulated mortality risk 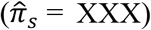 for NHB patients was 0.0069, reflecting a hypothetical absolute decrease in mortality rate of 0.69% (95% confidence interval (CI): (0.55%, 0.84%)). In our stratified analyses done by index quarter, results varied substantially. In Quarter 1, which included patients with index dates between March 1, 2020 and May 31, 2020, the average difference between observed mortality rate 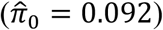 and simulated mortality risk was significantly greater at 0.0161, reflecting a hypothetical absolute decrease in mortality rate of 1.61% (95% CI: (1.51%, 1.70%)). The difference found in Quarter 2 (index dates between June 1, 2020 and August 31, 2020) was closer to the overall difference, with simulated mortality risk lower than observed mortality rate 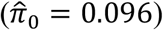 by 0.0051 on average (0.51% absolute difference, 95% CI: (0.35%, 0.67%)). No statistically significant difference in mortality rate was found for Quarter 3 (index dates between September 1, 2020 and November 30, 2020), with an average difference between observed mortality rate 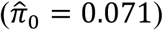 and simulated mortality risk of -0.0015 (−0.14% absolute difference, 95% CI: (−0.56%, 0.26%)). In Quarter 4 (index dates between December 1, 2020 and February 28, 2021), the difference between observed mortality rate 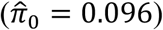 and simulated mortality risk was similar to that found in Quarter 1 with a mean difference of 0.0131 (1.31% absolute difference, 95% CI: (1.04%, 1.59%)). Differences between observed and simulated mortality rates across iterations varied most in Quarter 3 and least in Quarter 1.

**Figure 4.**
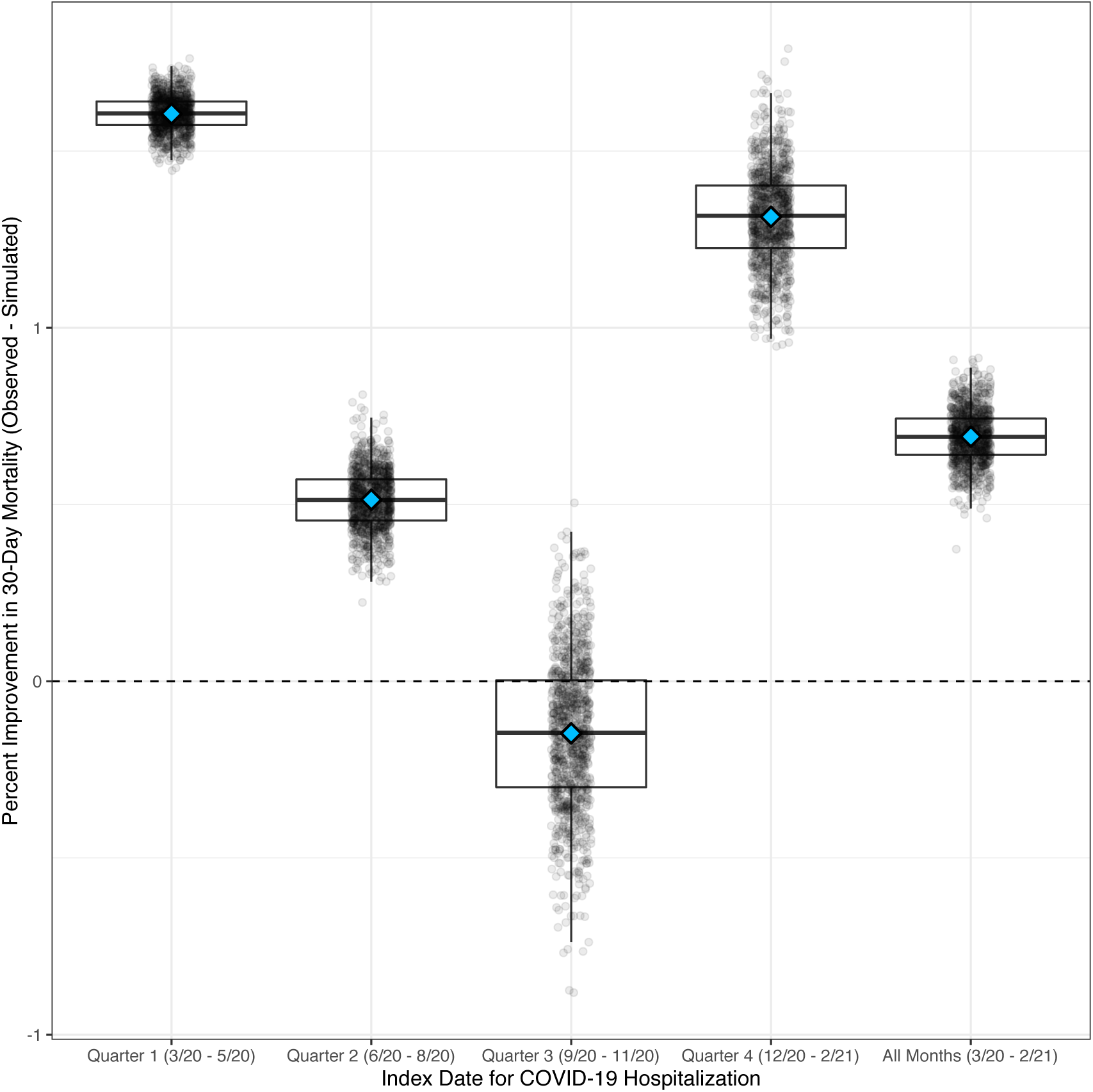
Boxplots depicting results from our five simulation studies, with index date for COVID-19 hospitalization on the *x*-axis (by index quarter and for all months) and the percent reduction in 30-day in-hospital mortality (observed mortality rate – simulated mortality risk) on the *y*-axis. For each boxplot, each circle represents the result from one replicate of the simulation. Blue diamonds in the center of each boxplot denote the mean difference across 500 replicates of the simulation.

## Discussion

In this work, we presented a novel application of a method for performing distributed generalized linear mixed modeling, the dPQL algorithm, to study the association between attending hospital and racial disparities in COVID-19 mortality. In real-world multi-site studies where hospitals are not able to share patient-level data with one another, this approach can be used to perform counterfactual modeling resulting in the exact same estimates that could be obtained if patient data were centralized, owing to the dPQL algorithm being lossless relative to traditional PQL. By not requiring patient-level data sharing, our proposed application could be used to study hospital-associated racial disparities using larger, more heterogeneous collections of patient data, allowing for more generalizable and clinically impactful conclusions. Our counterfactual modeling approach via simulation is also generalizable, able to be used in a variety of applied contexts beyond the application studied here. This approach could be used to investigate the association between any particular grouping of patients (e.g. at the hospital, state, or country level) and a non-continuous outcome of interest, with multinomial group assignments based on some exposure of interest.

Our real-world analysis of patient data from OneFlorida, while illustrative of our counterfactual modeling approach as a proof-of-concept analysis, is limited in several respects. Despite finding a slight improvement in mortality rate under the hypothetical scenario that NHB patients attend hospitals in the same distribution as NHW patients, we analyzed patient data from four Florida hospitals whose relative distributions of NHW and NHB patients did not differ substantially. The lack of patient heterogeneity among participating hospitals limited the ability of the GLMM fit by the dPQL algorithm to capture meaningful hospital-level differences with respect to in-hospital mortality risk. Our proposed application of the dPQL algorithm is best suited for real-world studies featuring large, heterogeneous collections of hospitals spanning a large geographic area, such as the study conducted in Asch et al. 2020 which included over 44,000 patients admitted to over 1,100 hospitals from 41 states [10]. Additionally, since our analysis was intended to be an illustrative example, there were no inclusion or exclusion criteria for covariates included in our fitted GLMM. In practice, if many candidate predictors are available, a variable section procedure should be used together with clinical expertise to select the final model and model fit should be assessed. Due to the limitations of our applied analysis, results presented should not be interpreted clinically. Rather, they are meant to illustrate the type of analysis that can be performed using our counterfactual modeling approach with multi-site data.

In future work, it would be worthwhile to compare different approaches for distributed generalized linear mixed modeling to counterfactually model in-hospital mortality. The dPQL method is one approach, but more are likely to become available in the near future. It could also be beneficial to continue to investigate whether the association between attending hospital and racial disparities in mortality has changed over time. Our limited analysis did not suggest a trend in either direction, but analysis of a more complete, heterogeneous collection of patient data would provide more convincing conclusions in this regard.

Despite the limitations of our own real-world analysis, we believe this novel application of the dPQL algorithm can be used by researchers as a tool for identifying hospital-level inequities in patient outcomes associated with race. In the event that inequities are present and thought to be related to quality of care, hospital-level interventions may be needed to help close gaps in performance between predominately white and predominately black hospitals. While this would likely take considerable time to accomplish, we hope our method can help to highlight underlying disparities and aid in the process of addressing systemic racism in healthcare.

## Data Availability

OneFlorida data can be requested at https://onefloridaconsortium.org/front-door/; Since OneFlorida data is a HIPAA limited data set, a data use agreement needs to be established with the OneFlorida network.

